# Racial Disparities in Opioid Overdoses, Treatment Access, and Healthcare Utilization: A Comprehensive Claims-Based Analysis, 2020–2024

**DOI:** 10.64898/2026.05.08.26352752

**Authors:** Anisha Pandey

## Abstract

**Purpose:** Opioid overdose deaths disproportionately affect racial and ethnic minority populations in the United States, yet claims-based evidence characterizing the multi-dimensional structure of these disparities across incidence, treatment access, costs, and insurance coverage remains limited.

**Methods:** We conducted a retrospective cross-sectional and longitudinal cohort analysis using the HealthVerity Launch Sample, a large administrative claims database. The study population comprised 3,675,823 patients across 5 racial groups enrolled between 2020 and 2024. Eight primary analyses were conducted, including age-sex standardized overdose rates, temporal disparity trends, medication-assisted treatment (MAT) receipt, naloxone access, pharmacy costs, insurance payer type, care setting, and multivariable logistic regression for overdose risk.

**Results:** Black patients had the highest age-sex standardized overdose rate (363.4 per 100,000; rate ratio [RR] = 1.27 vs. White), and those with opioid use disorder (OUD) received MAT at a rate 35% lower than White patients (19.8% vs. 30.7%; RR = 0.645), driven primarily by a buprenorphine access deficit. AIAN patients demonstrated consistent multi-dimensional disadvantage across naloxone access, MAT engagement, and pharmacy costs. After adjustment for payer type, age, and sex, all non-White groups showed lower adjusted odds of overdose than White patients (Black OR = 0.87; AIAN OR = 0.25), with Medicaid enrollment carrying 7.06 times the overdose odds of commercial insurance.

**Conclusion:** Insurance type is the dominant predictor of overdose risk, and the disproportionate Medicaid enrollment of Black patients is both a consequence of structural disadvantage and access disparities. Targeted interventions such as buprenorphine expansion in Medicaid and enhanced naloxone distribution are recommended.

## 1 Introduction

The opioid crisis remains one of the most severe public health emergencies in the United States, with opioid overdose mortality exceeding 80,000 annually in recent years [1]. While earlier phases of the epidemic disproportionately affected White, rural, and middle-aged populations, the rapid proliferation of illicitly manufactured fentanyl has profoundly altered the demographic distribution of overdose mortality [2]. Black Americans have experienced steep increases in overdose death rates since 2015, with Black men now experiencing overdose mortality rates comparable to or exceeding those of White men in several states [3], [4]. Despite this epidemiological shift, treatment access disparities have not resolved commensurately. Black individuals with OUD consistently receive buprenorphine at substantially lower rates than White individuals [5], [6]. Structural factors including insurance type, prescriber availability, and systemic biases in clinical decision-making have been posited as mechanisms [7].

Claims-based analyses offer an opportunity to examine these disparities at population scale, capturing not only diagnoses and treatment receipt but also payer type, care setting, and pharmacy utilization. However, prior claims-based studies have often been limited in scope, focusing on single dimensions of disparity or restricted geographic populations. The present study addresses this gap through a comprehensive, multi-dimensional claims-based analysis of racial disparities across the opioid care continuum from 2020 to 2024, using the HealthVerity Launch Sample — a large, longitudinal national administrative database.

## 2 Methods

### 2.1 Data Source

We analyzed the HealthVerity Launch Sample, a longitudinal administrative claims database comprising medical, pharmacy, and enrollment records from a nationally representative sample of insured individuals in the United States. The database links patient-level records across facilities, payers, and dispensing events, enabling longitudinal tracking of diagnoses, prescriptions, and healthcare utilization.

### 2.2 Study Population

The analytic population comprised 3,675,823 patients with known race, gender, and year of birth enrolled at any point between January 1, 2020, and December 31, 2024. Approximately 6.3% of enrolled patients (n = 245,462) were excluded due to unknown race/ethnicity. Five mutually exclusive racial groups were examined: White (n = 2,584,029; 65.9%), Black (n = 514,201; 13.1%), Asian (n = 233,543; 6.0%), AIAN (n = 202,085; 5.2%), and Other/Mixed (n = 141,965; 3.6%). Hispanic/Latino ethnicity was not analyzed due to 57.2% missing ethnicity data. Race was treated as a socially mediated variable reflecting structural and environmental exposures rather than a biological determinant.

### 2.3 Exposure & Outcome Definitions

Opioid overdose was defined as one or more ICD-10-CM T40.x diagnosis codes (63 codes, including T40.0–T40.4, T40.6) across accidental, intentional, assault, and undetermined intent categories. OUD was defined as one or more F11.x codes (19 codes). MAT receipt was identified from pharmacy claims using validated National Drug Code (NDC) codelists: buprenorphine/naloxone (84 NDCs), naltrexone (31 NDCs), and methadone (40 NDCs). Naloxone was identified using 54 NDCs for naloxone and branded Narcan formulations. Methadone dispensed at opioid treatment programs (OTPs) is not captured in pharmacy claims and therefore represents an undercount of methadone utilization. Multiple overdose claims within the same calendar year for the same patient were counted as a single episode for rate calculations.

**Table 1.** Baseline Characteristics of the Study Population by Race (2020–2024)

| Characteristic | White<br>(n = 2,584,029) | Black<br>(n = 514,201) | Asian<br>(n = 233,543) | AIAN<br>(n = 202,085) | Other/Mixed<br>(n = 141,965) |
| --- | --- | --- | --- | --- | --- |
| Age, mean (SD) | 40.8 (23.1) | 37.2 (23.2) | 37.0 (21.5) | 39.9 (21.4) | 31.4 (21.6) |
| 18–24 | 252,608 (9.8%) | 53,783 (10.5%) | 23,734 (10.2%) | 18,075 (8.9%) | 18,312 (12.9%) |
| 25–34 | 342,641 (13.3%) | 71,487 (13.9%) | 39,028 (16.7%) | 31,548 (15.6%) | 19,628 (13.8%) |
| 35–44 | 349,179 (13.5%) | 67,558 (13.1%) | 36,523 (15.6%) | 35,127 (17.4%) | 18,515 (13.0%) |
| 45–54 | 313,303 (12.1%) | 58,619 (11.4%) | 28,925 (12.4%) | 29,896 (14.8%) | 14,127 (10.0%) |
| 55–64 | 397,313 (15.4%) | 61,917 (12.0%) | 27,962 (12.0%) | 25,964 (12.8%) | 13,205 (9.3%) |
| 65+ | 426,757 (16.5%) | 73,249 (14.2%) | 26,780 (11.5%) | 27,031 (13.4%) | 11,690 (8.2%) |
| Sex, n (%) |  |  |  |  |  |
| Male | 1,227,976 (47.5%) | 221,194 (43.0%) | 110,694 (47.4%) | 93,488 (46.3%) | 71,849 (50.6%) |
| Female | 1,356,053 (52.5%) | 293,007 (57.0%) | 122,849 (52.6%) | 108,597 (53.7%) | 70,116 (49.4%) |
| Insurance, n (%) |  |  |  |  |  |
| Commercial | 1,930,001 (74.7%) | 276,143 (53.7%) | 171,211 (73.3%) | 157,093 (77.8%) | 90,408 (63.7%) |
| Medicaid | 332,385 (12.9%) | 169,853 (33.0%) | 42,313 (18.1%) | 30,529 (15.1%) | 43,317 (30.5%) |
| Medicare | 320,699 (12.4%) | 68,011 (13.2%) | 19,889 (8.5%) | 14,306 (7.1%) | 8,146 (5.7%) |
| Overdose, n (%) | 7,582 (0.29%) | 1,720 (0.33%) | 579 (0.25%) | 162 (0.08%) | 184 (0.13%) |
| OUD Diag., n (%) | 29,663 (1.15%) | 5,309 (1.03%) | 1,705 (0.73%) | 351 (0.17%) | 746 (0.53%) |
| Any MAT, n (%) | 9,111 (30.7%) | 1,052 (19.8%) | 571 (33.5%) | 80 (22.8%) | 268 (35.9%) |

Note: AIAN = American Indian/Alaska Native; MAT = medication-assisted treatment; OUD = opioid use disorder. Age and sex distributions to be populated from patient-level enrollment data. Percentages calculated within each racial group.

### 2.4 Statistical Analyses

Eight primary analyses were conducted: (1) age-sex standardized overdose rates by race using direct standardization against the full enrolled population; (2) annual overdose rate trends and disparity trajectories from 2020 to 2024; (3) MAT receipt rates among OUD patients by race with chi-square tests for group differences; (4) naloxone prescription access among overdose patients by race; (5) MAT and naloxone pharmacy costs by race (median and mean, with Kruskal-Wallis tests); (6) insurance payer type distribution among overdose patients by race; (7) care setting and place-of-service analysis; and (8) multivariable logistic regression for binary overdose outcome (n = 591,325) with race, age group, sex, and payer type as predictors, reporting adjusted odds ratios (aORs) with 95% confidence intervals (CIs). A sensitivity analysis restricting the primary analyses to 2020–2023 complete calendar years was also conducted to address potential 2024 claims processing lag. All analyses were performed using Apache Spark (PySpark) with downstream statistical modeling and visualization conducted in Python.

## 3 Results

### 3.1 Overdose Rates by Race

Age-sex standardized overdose rates per 100,000 enrolled patients were: Black, 363.4 (RR = 1.27); White, 286.2 (reference); Asian, 261.5 (RR = 0.91); Other/Mixed, 172.4 (RR = 0.60); and AIAN, 81.1 (RR = 0.28). The markedly low AIAN rate likely reflects severe undercapture in commercial claims rather than true epidemiologic protection, as it diverges sharply from national surveillance data.

**Table 2.** Age-Sex Standardized Opioid Overdose Rates per 100,000 Enrolled Patients by Race (2020–2024)

| Race | Enrolled (n) | Overdose (n) | Crude Rate per 100k | Std. Rate per 100k | RR Crude (vs White) | RR Std. (vs White) |
| --- | --- | --- | --- | --- | --- | --- |
| Black | 514,201 | 1,720 | 334.5 | 363.4 | 1.140 | 1.270 |
| White | 2,584,029 | 7,582 | 293.4 | 286.2 | 1.000 (ref) | 1.000 (ref) |
| Asian | 233,543 | 579 | 247.9 | 261.5 | 0.845 | 0.914 |
| Other/Mixed | 141,965 | 184 | 129.6 | 172.4 | 0.442 | 0.602 |
| AIAN | 202,085 | 162 | 80.2 | 81.1 | 0.273 | 0.283 |

### 3.2 Temporal Trends

The Black–White overdose rate disparity widened substantially during the study period, increasing from near parity in 2020 (RR = 1.04) to sustained elevation in 2022 (RR = 1.27) and 2023 (RR = 1.26). Black overdose rates increased at approximately 12 times the rate of White patients (+6.46 vs. +0.53 per 100,000 per year). Asian patients experienced a notable single-year rate increase of 40.5% in 2021.

**Fig. 1.**
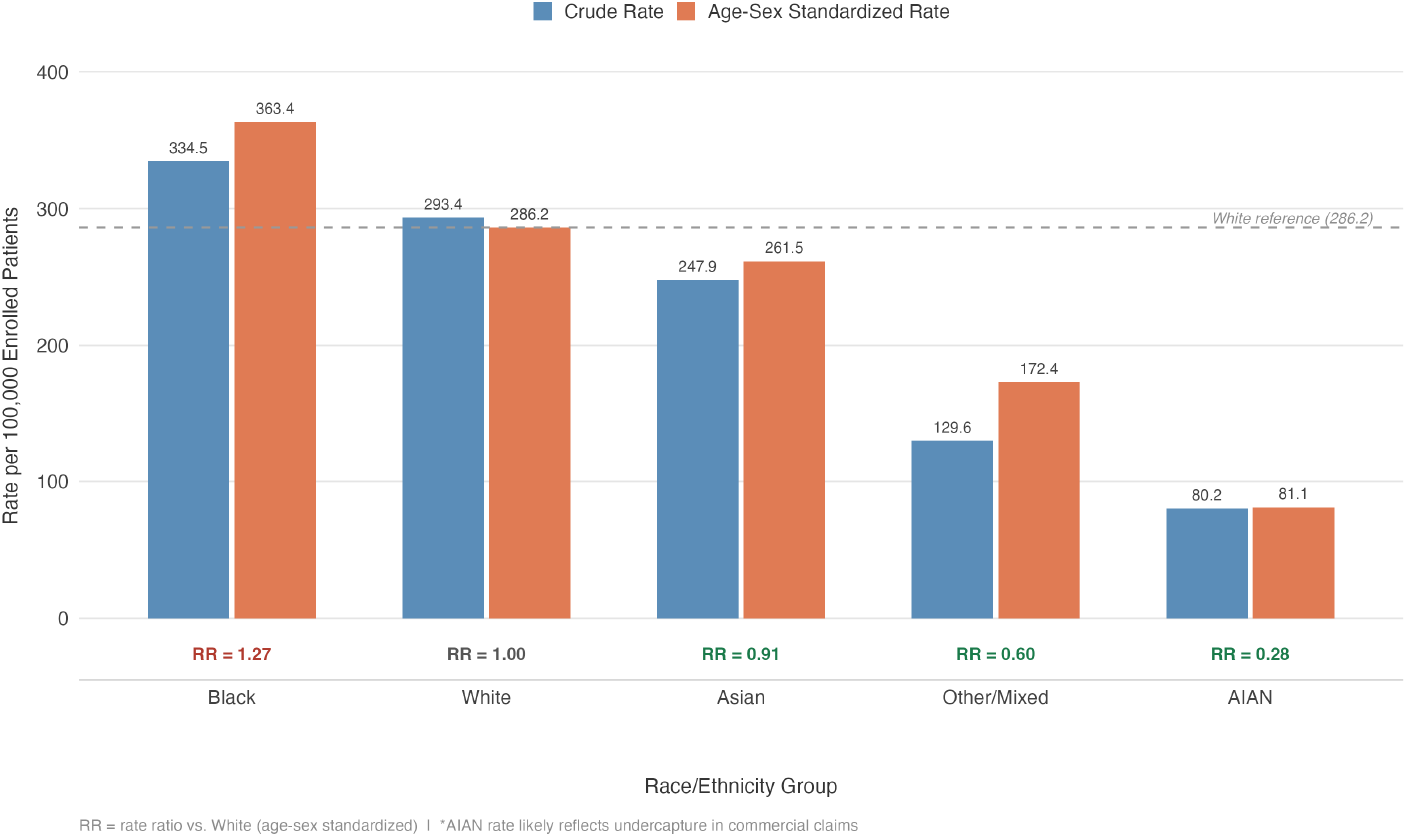
Opioid overdose rates per 100,000 patients by race. Blue bars represent crude rates; orange bars represent age-sex standardized rates. The dashed line indicates the White reference group (286.2).

**Table 3.** Annual Opioid Overdose Rates per 100,000 and Rate Ratios vs. White by Race (2020–2024)

| Race | 2020 | 2021 | 2022 | 2023 | 2024 |
| --- | --- | --- | --- | --- | --- |
| *Rate per 100,000* |  |  |  |  |  |
| Black | 71.6 | 81.9 | 92.2 | 89.7 | 60.9 |
| White | 68.7 | 73.8 | 72.4 | 70.9 | 52.6 |
| Asian | 47.5 | 66.8 | 65.9 | 60.4 | 45.0 |
| Other/Mixed | 27.5 | 26.1 | 37.3 | 32.4 | 20.4 |
| AIAN | 18.8 | 16.8 | 20.8 | 19.3 | 14.9 |
| *Rate Ratio vs. White* |  |  |  |  |  |
| Black | 1.042 | 1.109 | 1.273 | 1.265 | 1.157 |
| Asian | 0.692 | 0.905 | 0.911 | 0.852 | 0.854 |
| Other/Mixed | 0.400 | 0.353 | 0.516 | 0.457 | 0.388 |
| AIAN | 0.274 | 0.228 | 0.287 | 0.272 | 0.282 |

Note: 2024 rates are attenuated due to claims processing lag. RR = rate ratio vs. White reference.

### 3.3 MAT Treatment Disparities

Among 37,774 OUD patients, Black patients received any MAT at a rate of 19.8% compared with 30.7% for White patients (RR = 0.645; -10.9 percentage points). This deficit was driven almost entirely by lower buprenorphine access (17.5% vs. 27.6%; RR = 0.634). AIAN patients were also undertreated relative to White patients (any MAT RR = 0.743; -7.9 percentage points), with the deepest naltrexone gap (RR = 0.440). Asian (33.5%) and Other/Mixed (35.9%) patients received MAT at rates equal to or exceeding White patients.

### 3.4 Naloxone Access

Among overdose patients, AIAN patients had the lowest naloxone prescription rate (20.4% vs. 29.1% for White; RR = 0.70), as well as the lowest share with any MAT or naloxone pharmacy cost (24.1%) and the lowest median pharmacy cost ($23 vs. $124–$187 for other groups).

**Fig. 2.**
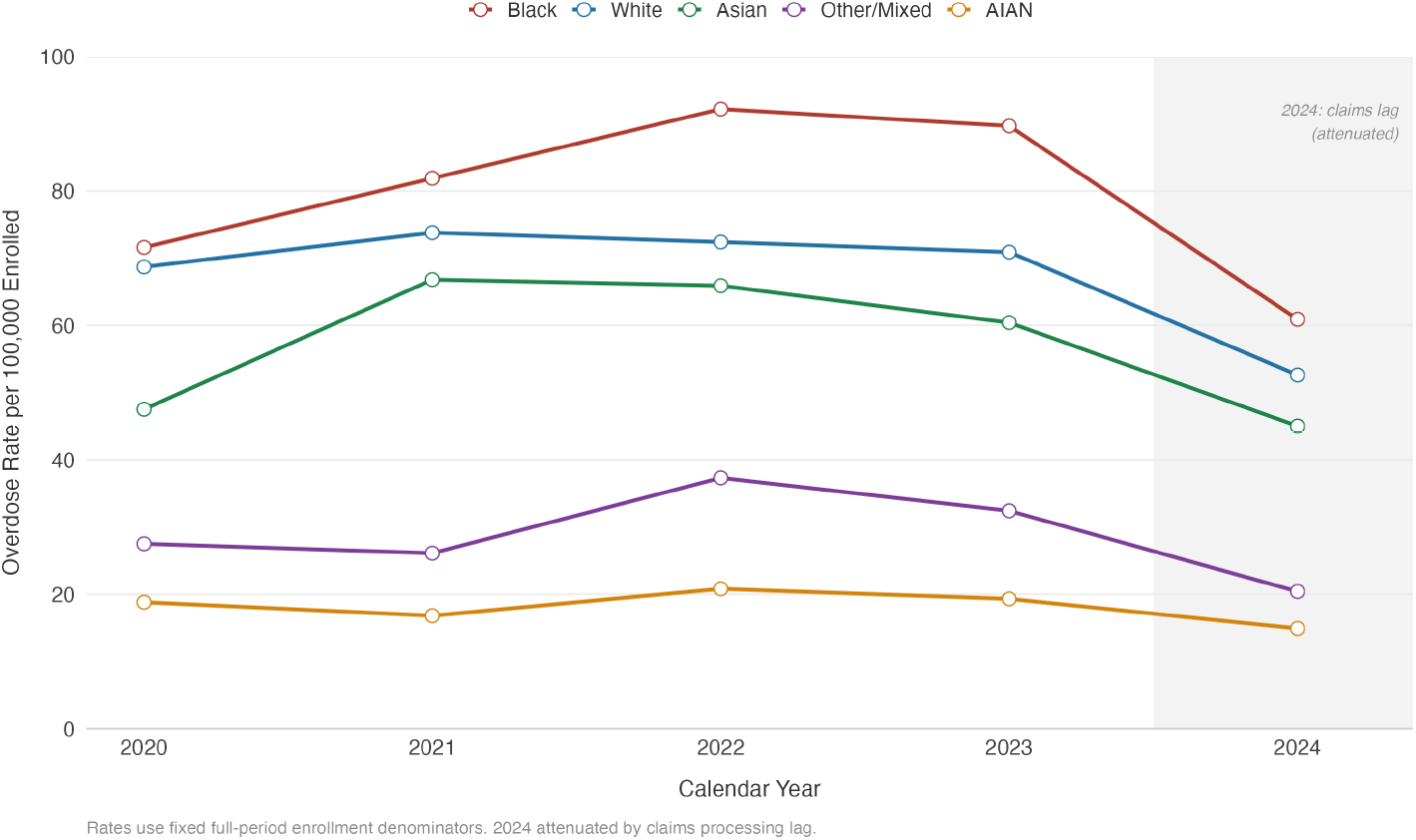
Annual opioid overdose rates per 100,000 enrolled patients by race, 2020–2024. Each line represents the annual overdose rate for one racial group. The shaded region indicates 2024, where rates are attenuated due to claims processing lag. AIAN = American Indian/Alaska Native.

**Table 4.** Medication-Assisted Treatment Receipt Rates by Race Among OUD Patients (2020–2024))

| Race | OUD<br>(n) | Buprenorphine<br>(n, %) | Naltrexone<br>(n, %) | Methadone<br>(n, %) | Any MAT<br>(n, %) |
| --- | --- | --- | --- | --- | --- |
| White | 29,663 | 8,187 (27.6) | 741 (2.5) | 504 (1.7) | 9,111 (30.7) |
| Black | 5,309 | 929 (17.5) | 96 (1.8) | 74 (1.4) | 1,052 (19.8) |
| Asian | 1,705 | 530 (31.1) | 27 (1.6) | 22 (1.3) | 571 (33.5) |
| AIAN | 351 | 71 (20.2) | 4 (1.1) | 7 (2.0) | 80 (22.8) |
| Other/Mixed | 746 | 237 (31.8) | 24 (3.2) | 16 (2.1) | 268 (35.9) |

Note: OUD = opioid use disorder; MAT = medication-assisted treatment; RR = rate ratio vs. White reference. Methadone figures are likely undercounted as opioid treatment program–dispensed methadone is not captured in pharmacy claims.

### 3.5 Insurance Type as a Structural Mediator

Payer type distribution differed substantially by race among overdose patients: Black and Other/Mixed patients were predominantly Medicaid-insured (51–52%), while White and Asian patients had more balanced commercial/Medicaid coverage (37% each). Multivariable logistic regression (n = 591,325; 9,920 overdose cases) identified payer type as the dominant predictor of overdose: Medicaid aOR = 7.06 (95% CI: 6.72–7.41); Medicare aOR = 4.63 (95% CI: 4.29–4.98), both relative to commercial insurance. Critically, after adjustment for payer type, age, and sex, all non-White racial groups demonstrated lower adjusted odds of overdose compared with White patients: Black aOR = 0.87; Asian aOR = 0.89; Other/Mixed aOR = 0.51; AIAN aOR = 0.25. This reversal indicates that insurance access and type functions as a major confounder and potential mediator of racial overdose disparities.

**Fig. 3.**
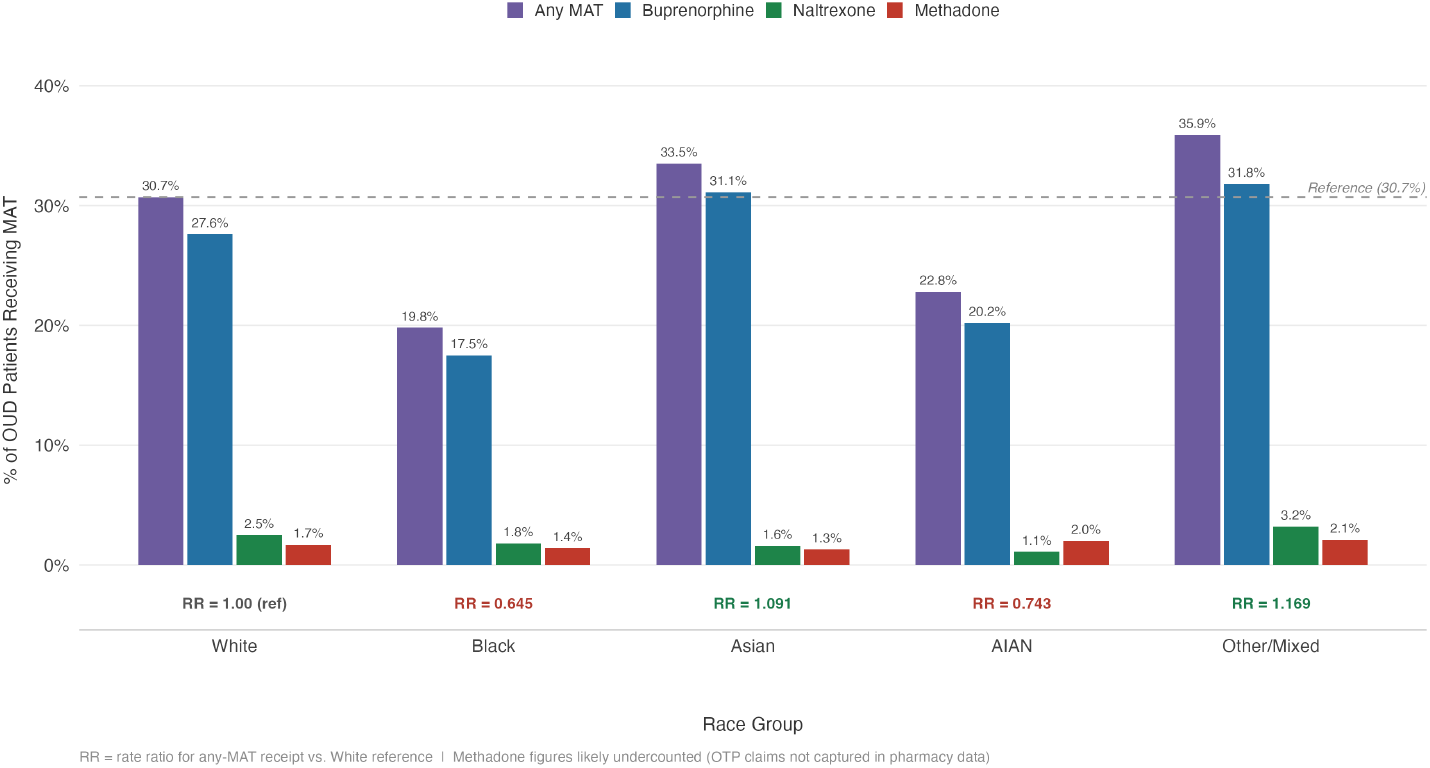
Medication-assisted treatment receipt rates by race among patients with opioid use disorder, 2020–2024. Grouped bars show the percentage of OUD patients receiving any MAT (purple), buprenorphine (blue), naltrexone (green), and methadone (red) within each racial group. The dashed reference line indicates the White any-MAT rate (30.7%). Rate ratios vs. White are displayed below each group. OUD = opioid use disorder; MAT = medication-assisted treatment.

**Table 5.** Naloxone Prescription Rates by Race Among Overdose Patients (2020–2024)

| Race | # of Overdoses (n) | # with Naloxone Rx (n) | Rate of Naloxone Rx (%) | RR |
| --- | --- | --- | --- | --- |
| White | 7,582 | 2,209 | 29.1 | 1.000 (ref) |
| Asian | 579 | 174 | 30.1 | 1.032 |
| Black | 1,720 | 508 | 29.5 | 1.014 |
| Other/Mixed | 184 | 52 | 28.3 | 0.970 |
| AIAN | 162 | 33 | 20.4 | 0.700 |

Note: RR = rate ratio vs. White reference. Naloxone prescriptions identified from pharmacy claims only; community distribution programs not captured.

### 3.6 Care Settings

Professional versus institutional claim-type distributions were uniform across racial groups (58% professional, 42% institutional). Exceptions included higher inpatient hospital utilization among Black patients (19.8% vs. 15.3% for White) and markedly lower telehealth utilization (1.0% vs. 2.6%), suggesting a potential digital access gap.

### 3.7 Sensitivity Analyses

Restricting analyses to 2020–2023 produced a maximum absolute change in RR of 0.022 across all 10 race-outcome combinations, confirming that findings were directionally and quantitatively robust to 2024 claims processing lag.

**Table 6.**
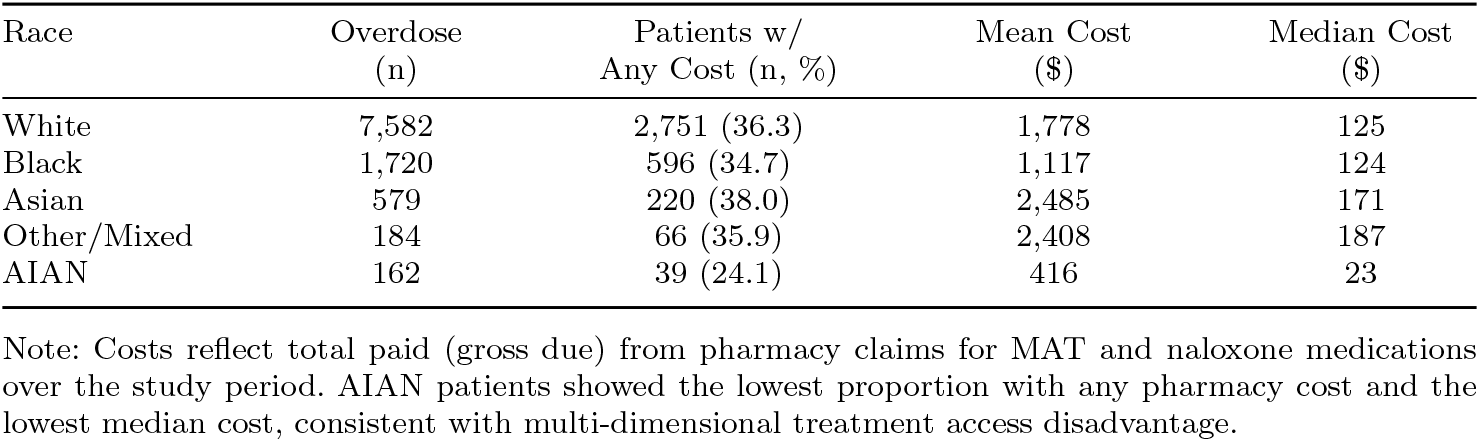
MAT and Naloxone Pharmacy Costs Among Overdose Patients by Race (2020–2024)

**Table 7.** Insurance Payer Type Distribution by Race Among Overdose Patients (2020–2024)

| Race | Commercial<br>(%) | Medicaid<br>(%) | Medicare<br>(%) | Other/Unspec.<br>(%) |
| --- | --- | --- | --- | --- |
| White | 36.1 | 37.3 | 25.9 | 0.7 |
| Black | 21.6 | 51.8 | 25.9 | 0.7 |
| Asian | 39.2 | 38.2 | 22.4 | 0.3 |
| Other/Mixed | 20.5 | 50.8 | 28.2 | 0.5 |
| AIAN | 36.7 | 44.4 | 18.7 | 0.3 |

Note: Payer type assigned based on most frequent payer across the study period. Black and Other/Mixed patients were predominantly Medicaid-insured ( 51%), compared with 37% for White patients.

**Table 8.**
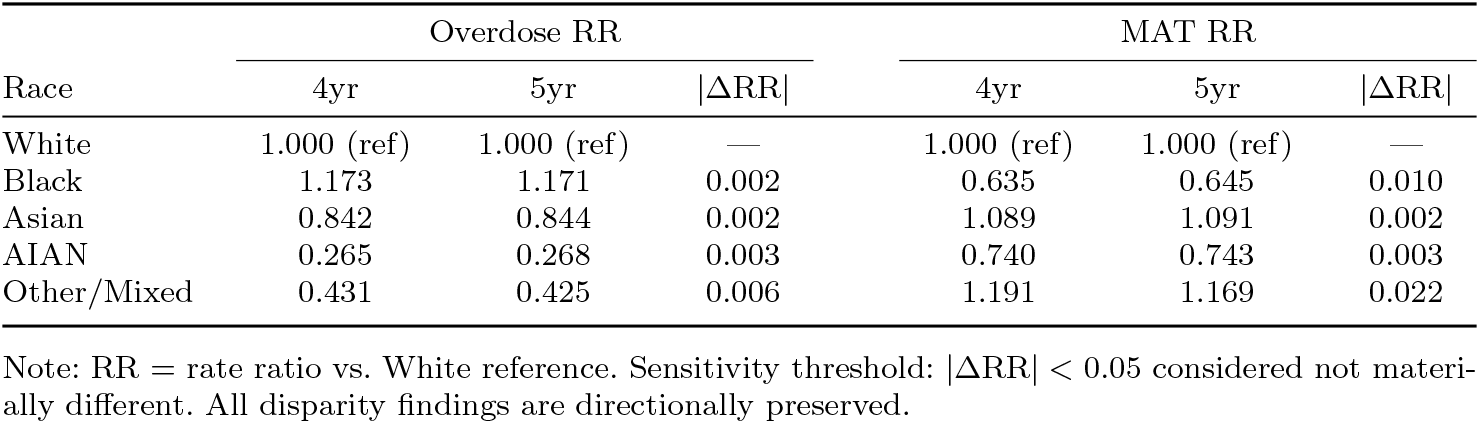
Sensitivity Analysis: Overdose Rate Ratios and MAT Treatment Rate Ratios — 4-Year (2020–2023) vs. 5-Year (2020–2024) Windows.

## 4 Discussion

This study reveals a multi-layered structure of racial disparities across the opioid care continuum. Black patients bear a disproportionate and widening overdose burden coupled with the largest MAT treatment access deficit among all racial groups, creating compounding disadvantage across incidence and treatment. The trajectory of the Black–White overdose disparity from near parity in 2020 to a 27% excess rate by 2022 is consistent with population-based surveillance data implicating the COVID-19 pandemic and accelerating fentanyl supply substitution as drivers of differential overdose risk [2], [8].

The finding that payer type dominates the adjusted prediction of overdose — with Medicaid enrollment carrying 7-fold higher odds than commercial insurance — reframes how racial disparities should be interpreted. The substantial reversal of race effects after payer adjustment suggests that much of the observed racial differential in overdose rates operates through the pathway of insurance-mediated barriers. These include Medicaid formulary restrictions on buprenorphine, step therapy and prior authorization requirements, lower reimbursement rates limiting buprenorphine-waivered provider availability in Medicaid networks, and clinic-level structural barriers concentrated in high-poverty, majority-Black communities [**?**], [**?**].

**Fig. 4.**
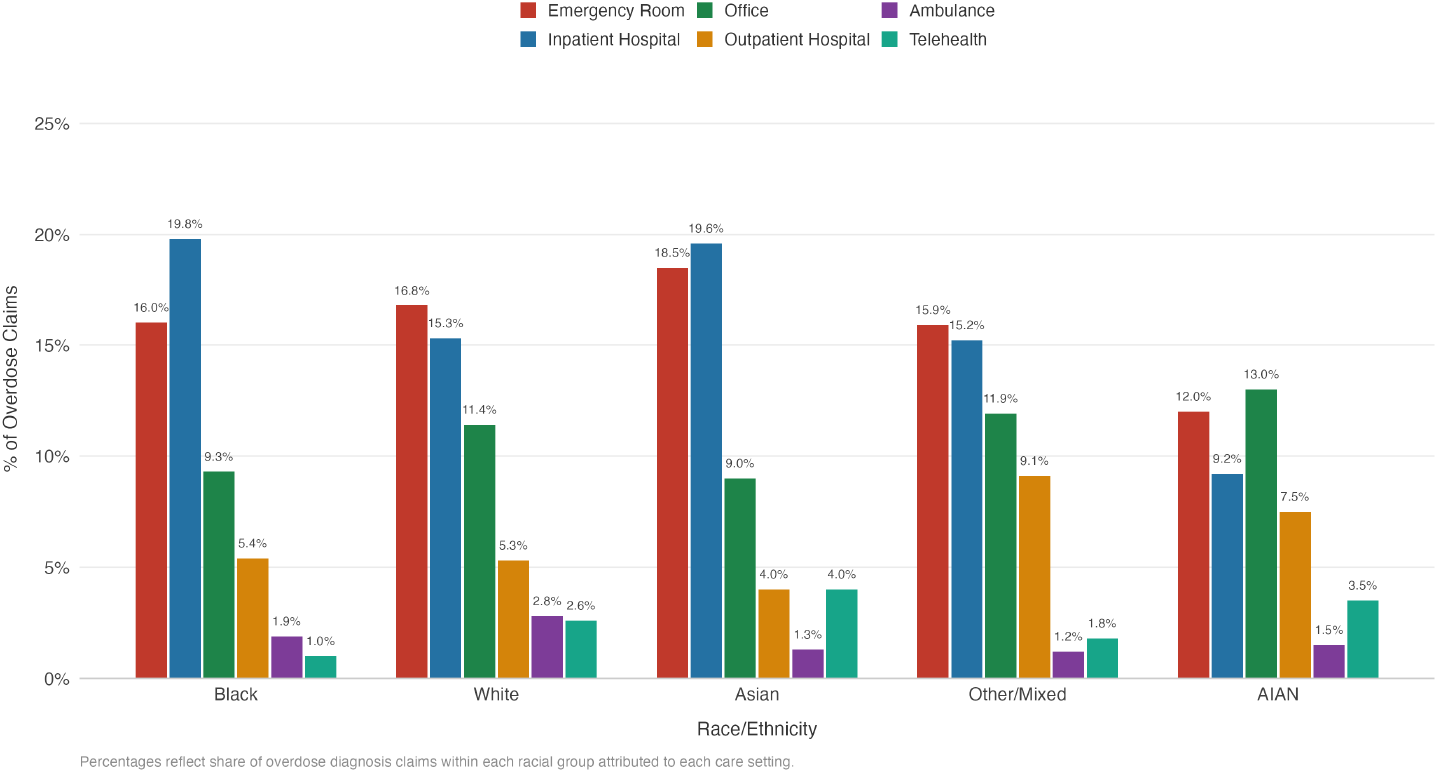
Place of service distribution for overdose-related claims by race, 2020–2024. Grouped bars show the percentage of overdose claims attributed to each of the six most common care settings within each racial group. Notable findings include higher inpatient hospital utilization among Black patients and lower telehealth utilization compared with White patients.

The treatment access deficit identified here — Black OUD patients receiving buprenorphine at 63% the rate of White patients — aligns with and extends prior work documenting racial disparities in buprenorphine initiation [5]. Addressing this deficit requires intervention at multiple levels, including expanding the prescribing workforce in Medicaid-dominated practicee, supporting culturally responsive OUD treatment programs, and eliminating prior authorization requirements [**?**], [**?**].

AIAN patients present a distinct challenge. While their claims-based overdose rates were paradoxically low, they demonstrated consistent disadvantage across naloxone access, MAT engagement, and pharmacy costs [4], [**?**]. Community-based naloxone distribution and tribally administered OUD treatment programs warrant dedicated investment and evaluation.

### 4.1 Limitations

This analysis is subject to several important limitations. First, results reflect an insured population and may not generalize to uninsured individuals, among whom overdose burden and access barriers may differ substantially. Second, AIAN patients are severely underrepresented in commercial claims; AIAN-specific findings should be interpreted with extreme caution and supplemented by data from tribal health systems and Indian Health Service records. Third, OTP-dispensed methadone is not captured in pharmacy claims, systematically undercounting methadone use — a limitation with disproportionate implications for Black patients, who use methadone at higher rates nationally.

Fourth, year-specific enrollment data was unavailable; period rates use total enrolled population as the fixed denominator across all years. Fifth, the cross-sectional payer assignment does not model payer transitions over the study period. Sixth, this is a descriptive and associational study; no causal claims are warranted, and adjusted analyses reduce but cannot eliminate residual confounding.

## 5 Conclusion

This comprehensive claims-based analysis documents a multi-dimensional structure of racial disparities in the opioid crisis: a widening Black–White overdose rate gap, a persistent 35% MAT access deficit for Black OUD patients, and consistent multi-dimensional disadvantage for AIAN patients. These findings are situated within a broader national context in which fentanyl-involved overdose deaths rose 100-fold among non-Hispanic Black individuals between 2010 and 2022, and in which structural racism has been identified as a fundamental contributor to drug overdose disparities, operating through interlocking mechanisms that span residential segregation, provider distribution, and insurance policy [9], [10].

The MAT access deficit documented here is consistent with and extends a growing body of national evidence. In the 180 days following an OUD-related index event, Black Medicare patients received buprenorphine after only 12.7% of events compared with 23.3% for White patients [11]. Substantial racial and ethnic disparities in the receipt of OUD medications exist especially between White and Black patients, despite similar health care utilization across racial and ethnic groups — suggesting that differential access, not differential need or healthcare contact, is the operative mechanism [11]. Despite overall improvements in MOUD access over time, significant racial and insurance-based disparities persist and require urgent, targeted solutions [12].

Insurance type — particularly Medicaid enrollment — is the dominant structural predictor of overdose risk, and the concentration of Black patients in Medicaid perpetuates treatment access barriers as shown in our adjusted models. Policy interventions must therefore be multipronged. The evidence on prior authorization removal is nuanced: removal of Medicaid prior authorization for buprenorphine was associated with increased prescribing in states with low baseline prescribing, but not in states with higher baseline prescribing, indicating that PA elimination alone is insufficient without concurrent workforce and capacity investment [13]. Expansion of the buprenorphine-prescribing workforce in Medicaid-dominant settings, culturally responsive OUD treatment programs to address provider bias and medical mistrust, community-based naloxone distribution for AIAN communities, and telehealth infrastructure investment for Black patients with OUD represent complementary and necessary levers [9], [10], [12].

Addressing the overdose crisis and racial disparities in addiction will likely require community-specific interventions that engage with minority populations and the clinicians who serve them to reduce stigma and bolster trust. The reversal of race effects after payer adjustment in our models underscores that insurance-mediated structural barriers — not race as a biological attribute — are the proximate drivers of disparity. Achieving health equity in the opioid crisis demands policy action commensurate with the structural depth of these inequities.

## Data Availability

All data produced in the present study are available upon reasonable request to the authors.

## Notes

### Competing Interest Statement

The authors have declared no competing interest.

### Funding Statement

This study did not receive any funding

### Author Declarations

The analyses in this study were conducted using individual-level patient data from the HealthVerity Launch Sample, a commercially licensed administrative claims database. All individual-level data were de-identified by HealthVerity prior to access and use, in accordance with HIPAA Safe Harbor standards. No re-identification of patients was attempted or possible in the course of this research.

